# Identifying and alleviating bias due to differential depletion of susceptible people in post-marketing evaluations of COVID-19 vaccines

**DOI:** 10.1101/2021.07.15.21260595

**Authors:** Rebecca Kahn, Stephanie J. Schrag, Jennifer R. Verani, Marc Lipsitch

**Affiliations:** Center for Communicable Disease Dynamics, Department of Epidemiology, Harvard T.H. Chan School of Public Health, Boston, Massachusetts; COVID-19 Response, US Centers for Disease Control and Prevention, Atlanta, Georgia; Department of Immunology and Infectious Diseases, Harvard T.H. Chan School of Public Health, Boston, Massachusetts

## Abstract

Recent studies have provided key information about SARS-CoV-2 vaccines’ efficacy and effectiveness (VE). One important question that remains is whether the protection conferred by vaccines wanes over time. However, estimates over time are subject to bias from differential depletion of susceptibles between vaccinated and unvaccinated groups. Here we examine the extent to which biases occur under different scenarios and assess whether serologic testing has the potential to correct this bias. By identifying non-vaccine antibodies, these tests could identify individuals with prior infection. We find in scenarios with high baseline VE, differential depletion of susceptibles creates minimal bias in VE estimates, suggesting that any observed declines are likely not due to spurious waning alone. However, if baseline VE is lower, the bias for leaky vaccines (that reduce individual probability of infection given contact) is larger and should be corrected by excluding individuals with past infection if the mechanism is known to be leaky. Conducting analyses both unadjusted and adjusted for past infection could give lower and upper bounds for the true VE. Studies of VE should therefore enroll individuals regardless of prior infection history but also collect information, ideally through serologic testing, on this critical variable.

---

Vaccines are a critical tool for combatting the COVID-19 pandemic. Clinical trials and observational studies have provided key information about the vaccines’ efficacy and effectiveness (VE). One important question that remains to be answered is whether or not the protection conferred by vaccines wanes over time. However, estimates of effectiveness over time are subject to bias from differential depletion of susceptibles between vaccinated and unvaccinated groups. This bias occurs when individuals who are no longer at risk of infection due to protection from past infection are included in the analysis; assuming the VE is greater than zero, these individuals with prior infection are more likely to be unvaccinated than vaccinated. Therefore, over time, more uninfected and unvaccinated individuals who are not at risk of infection are included in the analysis, biasing VE estimates downward. This bias grows as infection spreads and makes the VE incorrectly appear to wane over time (i.e. spurious waning) (1–4). Although some studies attempt to restrict analysis to those without prior infection, often many past infections will go undetected or unreported, particularly for pathogens with a large proportion of asymptomatic or mild infections. Additionally, in a population with individuals who have heterogeneous risk of infection (for example due to occupational exposure or choice to wear a face covering), the riskiest individuals will be depleted preferentially among the unvaccinated group when the vaccine is effective, leading to the same bias downwards in VE, growing over time and thus seemingly showing waning of VE (1).

Serologic testing for SARS-CoV-2 antibodies has the potential to help correct the first bias. By identifying non-vaccine antibodies (e.g. N-protein), these tests could be used to identify individuals with prior infection and exclude them from studies of VE over time. Likewise, adjustment for individual-level risk of infection (in practice, for proxies such as occupation or behavior) can help address the second bias.

While each of these issues can in principle affect VE estimates and induce a spurious impression of waning VE, the magnitude of this bias under various assumptions about baseline VE is not clear, nor has it been shown before to our knowledge how adjustments can solve the problems. Here we examine the extent to which these biases occur under different scenarios and assess approaches to alleviate bias under various assumptions.

## Methods

### Network and epidemic

We first create a network model of 20,000 individuals, similar to models described previously (2,5). The probability of connections between individuals in the network is calibrated in combination with the parameter for the probability of infection given contact to result in a reproduction number (R) of 1.25 or 1.50 (see Table 1 for a full list of parameters) (6). We seed an epidemic of a SARS-CoV-2-like pathogen with ten exposed individuals. Each day, each susceptible individual has a daily probability of infection from their infected connections in the network. A random half of the population is high risk, and the other half is low risk. High risk individuals have a daily probability of infection three times that of low risk individuals. This binary risk status is a simplified proxy for multiple factors that could affect individuals’ risks for infection, such as occupation, demographics, geography, or behavioral patterns (7–9).

**Table 1.** Parameters and associated values used in network model simulations

| **Parameter** | **Value(s)** |
| --- | --- |
| Population size | 20,000 |
| Average degree (number of connections in network) | 36, 43, 58 |
| Average beta (probability of infection per contact) | 0.005 |
| Reproduction number (R) | 1.25, 1.50, 2.00 |
| Proportion symptomatic | 0.5, 0.8 (33) |
| Number of initial cases | 10, 20 |
| Incubation period (days) | 5 (34) |
| Infectious period (days) | 7 |
| Proportion at high risk | 0.5, 0.1 |
| Relative beta for high risk | 1.5, 3.57 |
| Relative beta for low risk | 0.5, 0.71 |
| Days with complete protection from past infection | 90 (10) |
| Relative beta for those recovered after 90 days | 0.05, 0.3 (11,24) |
| Proportion of population vaccinated | 0.125 |
| Proportion previously recovered at start of simulation | 0, 0.1, 0.2, 0.3 |

We assume that half of those who are infected become symptomatic and that people are infectious for seven days. We assume that symptomatic, pre-symptomatic, and asymptomatic infected individuals have the same level of infectiousness. After individuals recover, we assume that complete protection from natural immunity lasts for 90 days (10), after which individuals can be reinfected; we then assume recovered individuals’ susceptibility is 95% lower than those without prior infection, resulting in low numbers of reinfection during the study period examined in the simulations (11). It is unknown exactly how VE differs for recovered individuals, although there is evidence that vaccination further reduces previously infected individuals’ risk (12). For simplicity we assume vaccinated recovered individuals’ susceptibility is further decreased by the same amount as for vaccinated susceptible individuals.

### Scenarios

We simulate random vaccination (to prevent unmeasured confounding) of 2500 individuals, or 12.5% of the population, on the first day of the simulation. Another 2500 unvaccinated individuals are also randomly selected for potential follow-up over the course of the simulations. We compare four primary scenarios (Table 2). In the first scenario, vaccine efficacy against susceptibility to infection (VE_S_) is 0.90, and vaccine efficacy against progression to symptoms (VE_P_) is 0.5. These measures combine to give a vaccine efficacy against symptomatic disease (VE_SP_), the primary outcome of most SARS-CoV-2 vaccine trials (13–16), of 0.95, under the formula *VE*_*SP*_ *= 1* − *(1* − *VE*_*S*_*)(1* − *VE*_*P*_)(17). These values are similar to those that have been observed in the trials (13,15) and initial observational studies (18–20) of the mRNA vaccines. In the second scenario, we assume VE_S_ and VE_SP_ are 0.7, similar to the findings from the Janssen vaccine trial (14).

**Table 2.**
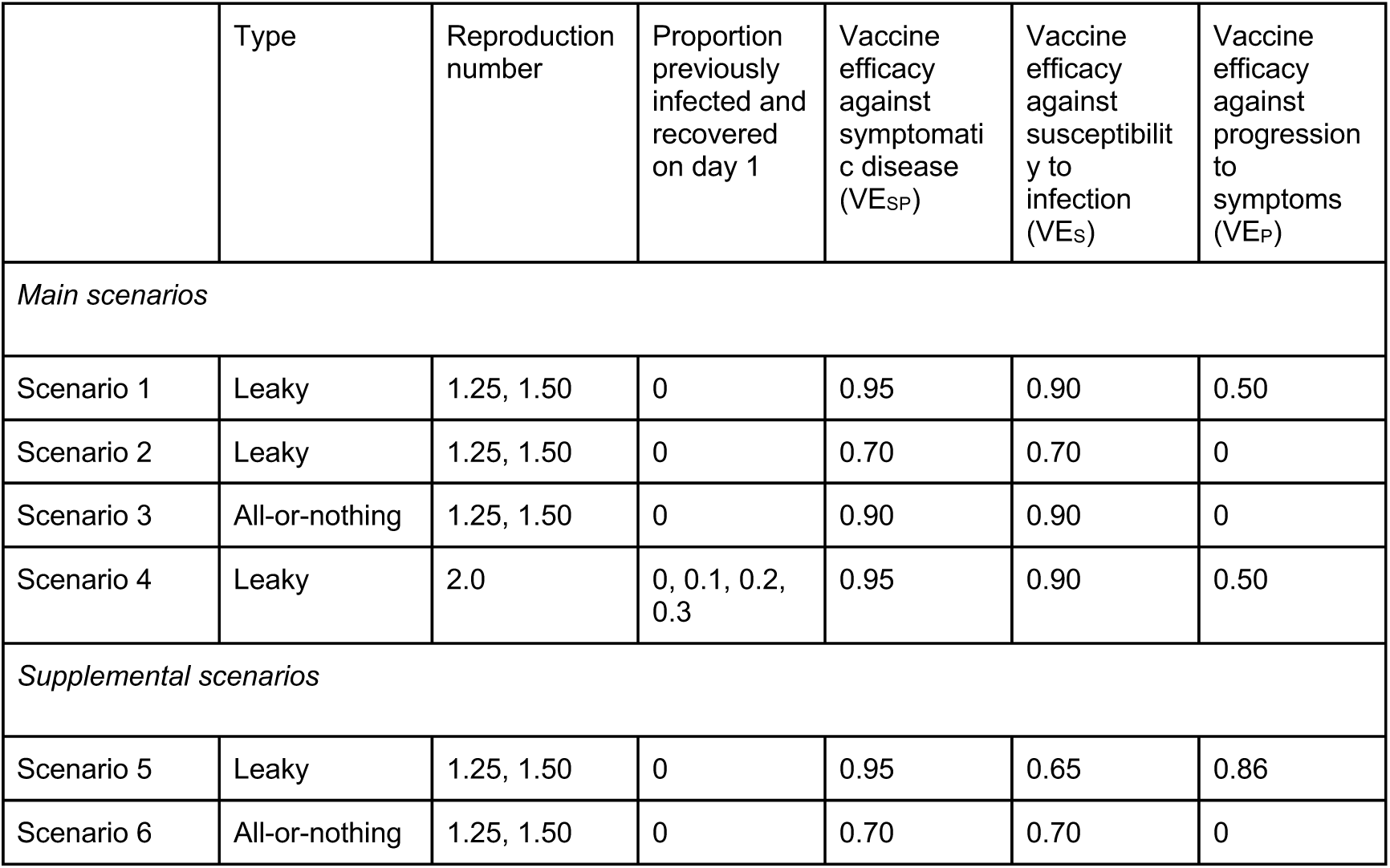
Scenarios the network model evaluated to assess the potential for spurious waning

In the first two scenarios, we assume the vaccine is “leaky”, meaning it reduces the probability of infection given contact to an equal degree, but not perfectly, in all vaccinated individuals (3). However, in the third scenario, to assess the impact of the vaccine mechanism, we model an all-or-nothing vaccine, meaning it protects a certain proportion of vaccinated individuals completely and provides no protection to the rest. In this scenario, VE_S_ and VE_SP_ are both 0.9. In supplementary scenarios, we also examine an all-or-nothing vaccine with lower VE_S_ and VE_SP,_as well as a leaky vaccine with VE_SP_ = 0.95, similar to Scenario 1, but with lower VE_S_ and higher VE_P._ (21).

Finally, in scenario 4, we examine a setting with a leaky vaccine with VE_SP_ = 0.95 in which some of the population has already been infected and recovered before the simulations and vaccination begin. We explore a range from 0-30% of individuals with prior infection under a higher R than in the other scenarios (R=2.0) to prevent herd immunity from prior infections from substantially slowing the epidemics before spurious waning can be observed. In these simulations, 20 individuals are exposed on the first day and 100 individuals are infectious (except in the simulations with 0 individuals previously infected).

### Analyses

#### Test-negative design

We then simulate sampling of cases, or individuals with COVID-19 (symptoms and positive virologic test), on a given day and a random 1:4 sample of controls (i.e. individuals without COVID-19), similar to a test-negative design (TND). We repeat this sampling for seven different time periods, every 25 days from day 75 to day 225, treating each day independently. Given the faster epidemics in scenario 4 with the higher R, we examine every 25 days from day 25 to day 150. We then estimate VE_SP_ -- the estimand that is in practice estimated in a standard TND, although the progression to symptoms aspect is not always acknowledged (22) -- using four analyses. We focus on VE_SP_ as it was the primary outcome in vaccine trials and due to potential biases that can arise in TNDs when estimating VE against all infection when vaccines affect disease severity (23).

In the first analysis (baseline), we estimate VE_SP_ by calculating the odds ratio (OR), using data from all individuals sampled:

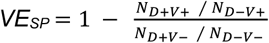, where D is disease (symptoms and a positive virologic test) and V is vaccine.

In the second analysis, we estimate the OR using logistic regression, controlling for risk (i.e. the binary measure described above for increased or decreased susceptibility to infection).

In the third analysis, we simulate serologic testing for non-vaccine antibodies (i.e. evidence of past infection) and then restrict the analysis to individuals who had not previously been infected. In the fourth analysis, we both restrict to those without evidence of previous infection and also control for risk. In the primary analyses, we assume perfect sensitivity and specificity of the serologic test for prior infection, but we relax these assumptions in sensitivity analyses. We examine lower sensitivity for cases and controls and lower specificity for cases only, as antibodies detected could reflect either current or prior infection.

#### Cohort / randomized controlled trial design

As a comparison to the TND, we repeat the same four analyses to estimate VE using a cohort design, where the time of symptomatic infection is known for the 5000 people under follow-up. We again examine different lengths of follow-up for this study design. We assume no unmeasured confounding: that is, no common causes of vaccination and infection, as would be true with adequate control for confounders. In practice, this study could be done using an electronic health records database using stratification, matching, or modeling for example to control for confounding factors such as occupation, age, insurance, and other factors affecting both vaccination and the likelihood of infection given vaccination. Because there is no unmeasured confounding and vaccination is random in these simulations, this design is comparable to a randomized controlled trial (RCT) in which all symptomatic cases are identified.

#### Sensitivity analyses

We vary key parameters of interest to examine their impact on the results. First, we vary the reduction in susceptibility conferred by past infection, using a value of 70% (24) reduction compared to the baseline parameter of 95% reduction. Next, we vary the proportion of the population that is high risk, examining a scenario in which only 10% of the population is high risk, with five times higher risk than lower risk individuals. Finally, we vary the proportion of infections that are symptomatic, using a higher value of 0.8 (23) compared to the baseline of 0.5.

Ethics: This activity was reviewed by CDC and was conducted consistent with applicable federal law and CDC policy.

## Results

In scenario 1 with high VE_S_ and VE_SP_, we find that for most time points, all four TND analyses return estimates of VE_SP_ close to the true value of 0.95 (Figure 1). However, in the simulations with R=1.5, the first two analyses that do not exclude prior infection result in downward biases for days further from vaccination (i.e. as low as 0.89 200 days from vaccination). This bias occurs at later dates and in the higher transmission scenarios (i.e. when more cases have occurred) due to differential depletion of susceptibles between vaccinated and unvaccinated individuals over time; the bias is alleviated by excluding those with prior infection from the analysis. Similar results are found in an additional analysis with the same VE_SP_ but different VE_S_ and VE_P_ (Figure S1). Note, in these and other simulations with high VE, when the number of cases is very low at either the beginning or end of the epidemic, imprecision can result in VE estimates of 1 (if all cases are by chance unvaccinated).

**Figure 1.**
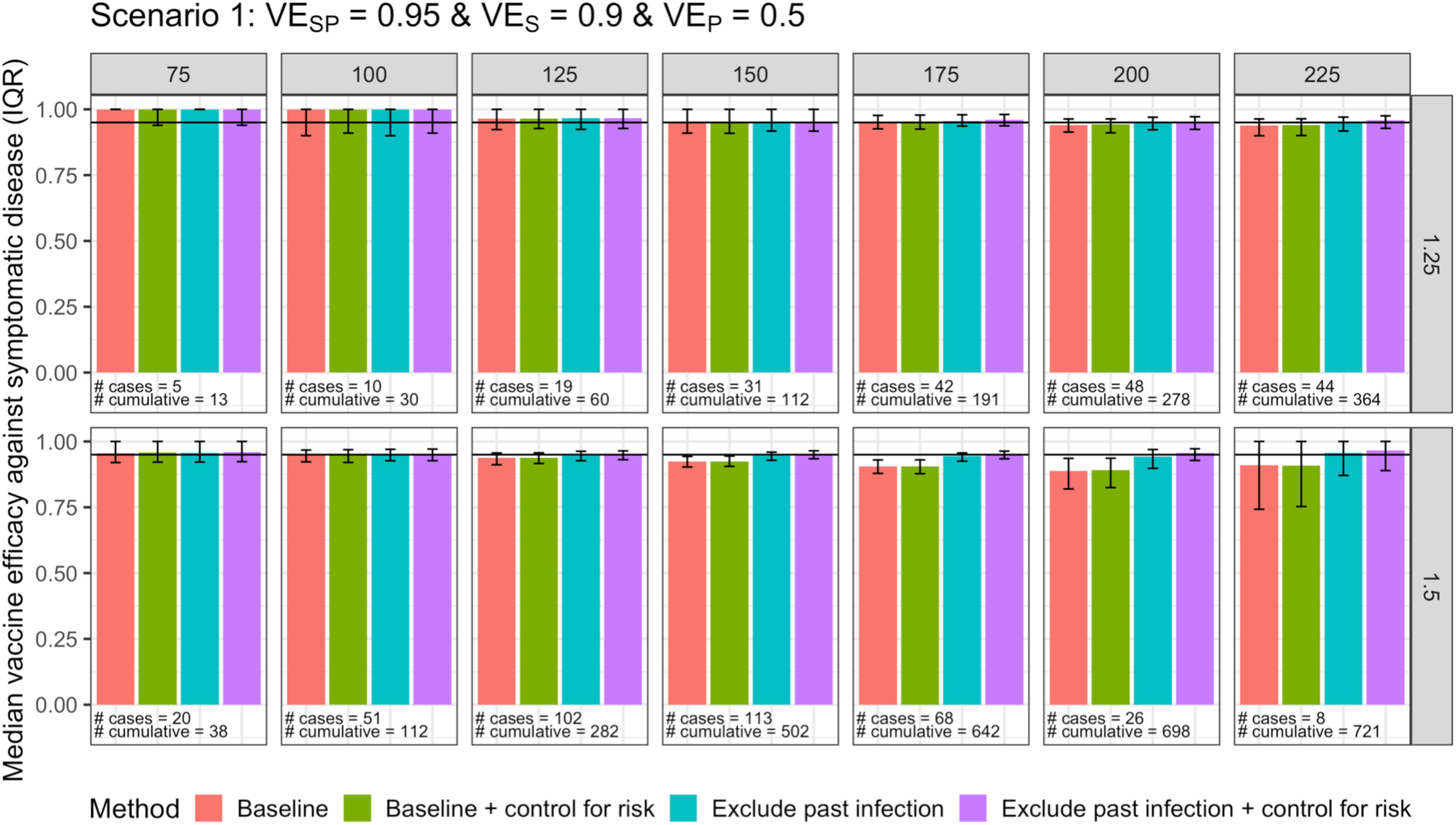
Vaccine efficacy against symptomatic disease for scenario 1 with a test-negative design. Columns are days since vaccination, and rows are values of R. Median and IQR of 100 simulations shown. Number of cases refers to the median number of people with COVID-19 included in that day’s analysis. Cumulative number refers to the median total number of cases of COVID-19 by that day since vaccination (denominator 5000).

In scenario 2, with a lower value of VE_S_ and VE_SP_ of 0.7, the first and second analyses that do not exclude prior infection are biased further downwards than in scenario 1 (i.e. as low as 0.36 225 days from vaccination when R = 1.5); this bias also occurs earlier than in scenario 1 and for both R values (Figure 2) because the epidemic is larger due to lower VE. In addition, in this scenario, the third analysis that excludes those with prior infection but does not control for risk has a more pronounced bias (lowest value of 0.64 compared to true VE_SP_ of 0.7 on day 225 when R=1.5) than in scenario 1 (lowest value of 0.94 compared to true VE_SP_ of 0.95 on day 200 when R=1.5).

**Figure 2.**
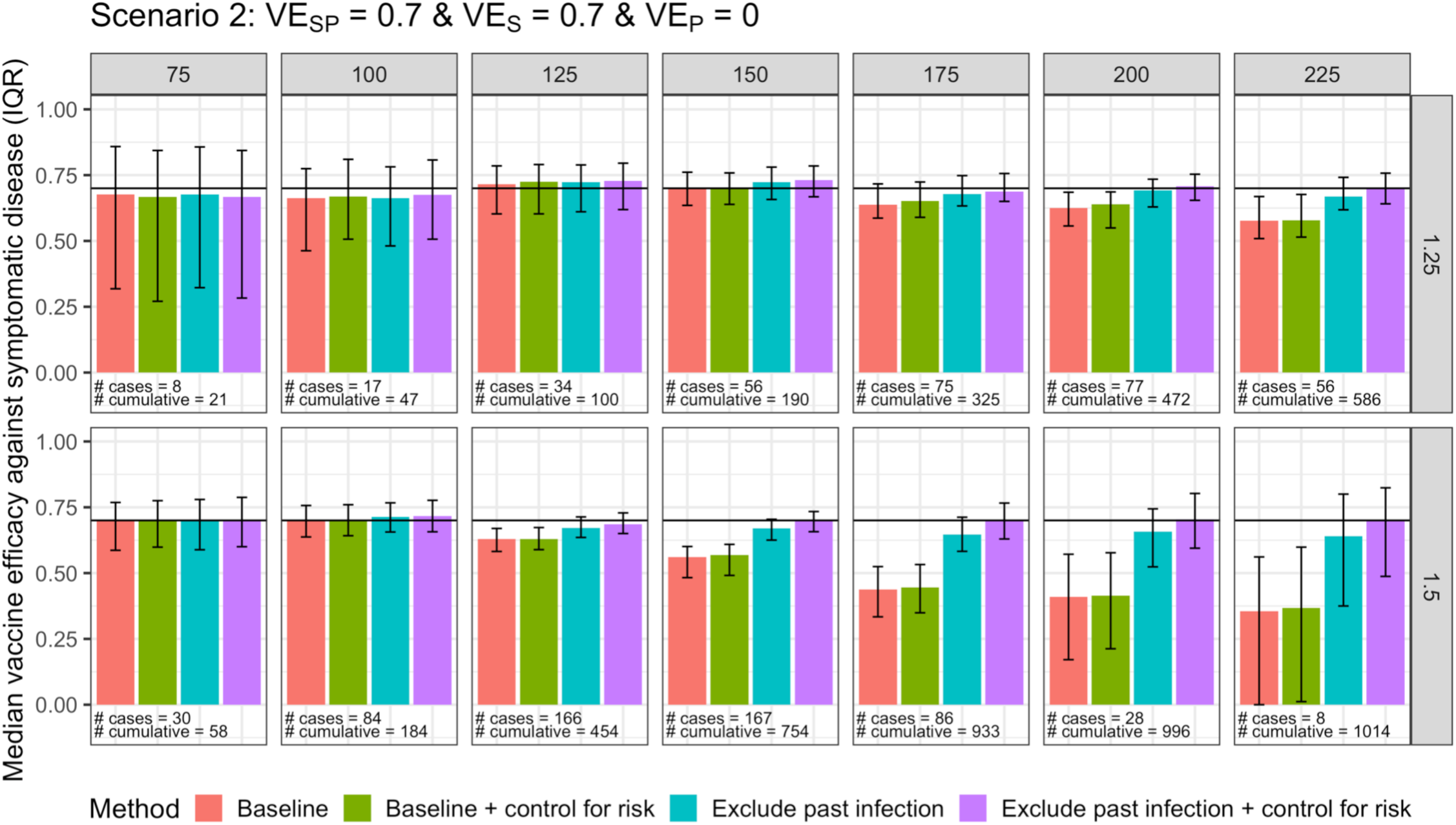
Vaccine efficacy against symptomatic disease for scenario 2 with a test-negative design. Columns are days since vaccination, and rows are values of R. Median and IQR of 100 simulations shown. Number of cases refers to the median number of people with COVID-19 included in that day’s analysis. Cumulative number refers to the median total number of cases of COVID-19 by that day since vaccination (denominator 5000).

In scenario 3, which models an all-or-nothing vaccine mechanism instead of a leaky mechanism, excluding prior infections results in a bias upward away from the true VE_SP_ of 0.9 (Figure 3), with some values approaching 1. This bias is more pronounced in the higher R simulations, on later days, and for lower values of VE_SP_; for example, on day 200 in the R=1.5 simulations, the VE_SP_ from the analysis excluding prior infection and adjusting for risk is 0.84, compared to the true value of 0.7 (Figure S2)

**Figure 3.**
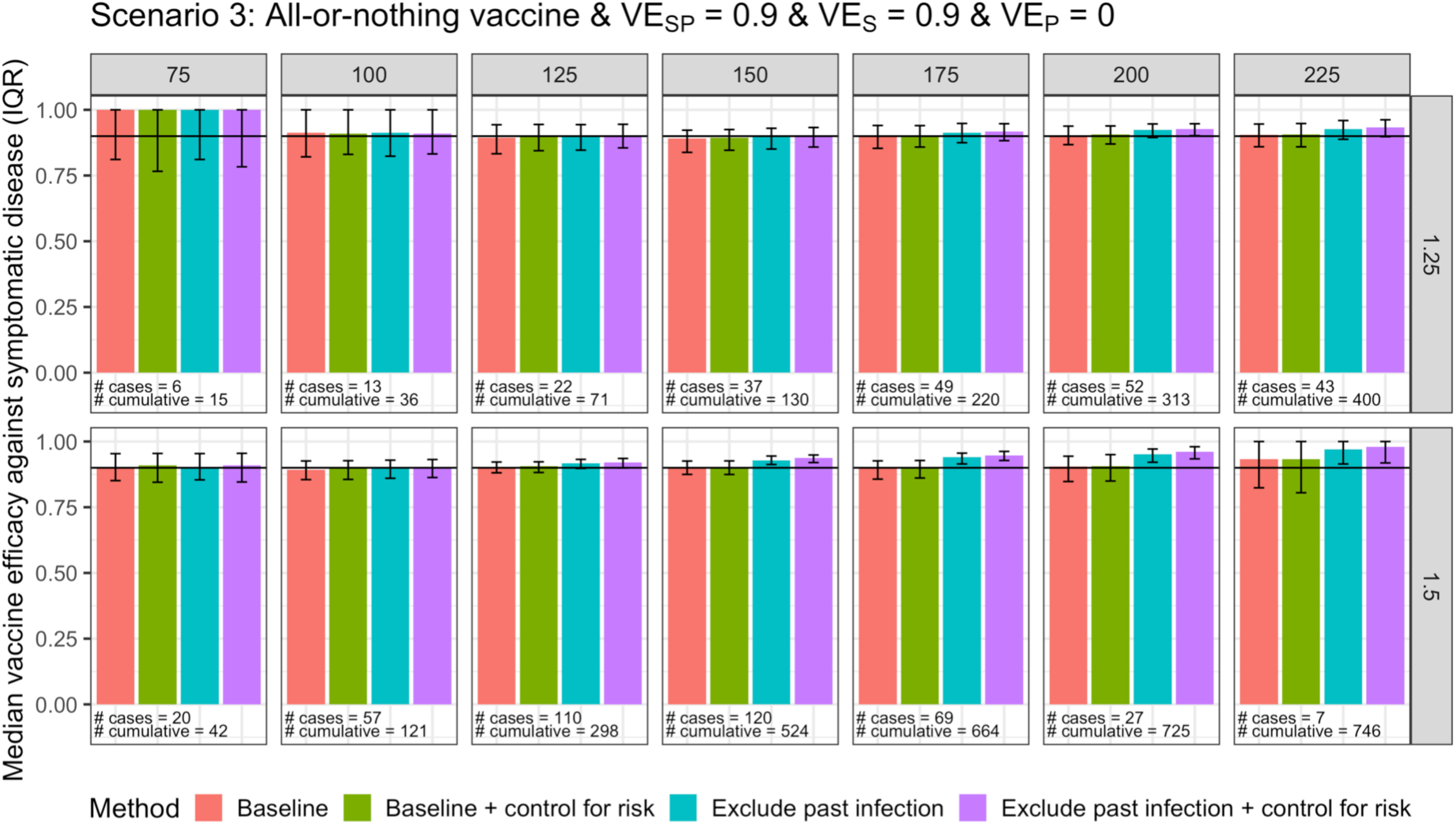
Vaccine efficacy against symptomatic disease for scenario 3 with a test-negative design. Columns are days since vaccination, and rows are values of R. Median and IQR of 100 simulations shown. Number of cases refers to the median number of people with COVID-19 included in that day’s analysis. Cumulative number refers to the median total number of cases of COVID-19 by that day since vaccination (denominator 5000).

In scenario 4, we see that the degree of spurious waning bias increases with the number of cumulative cases *since vaccination* (Figure 4). This trend occurs because the bias is driven by differential depletion of susceptibles between vaccinated and unvaccinated individuals. In the simulations with 0% prior infection at the time of vaccination, the epidemic and vaccination begin simultaneously; thus, when evaluating VE at later dates, because the vaccine reduces risk of infection, those who have been infected prior to the date of interest are more likely to be unvaccinated, causing bias. Assuming prior infection doesn’t affect the decision to be vaccinated, in the simulations where the epidemic begins prior to vaccination, the distribution of vaccination status among those infected becomes more balanced because the infections prior to vaccination are expected to be evenly split between vaccinated and unvaccinated individuals. This is why the bias increases with cumulative cases since vaccination began, rather than with overall cumulative cases (i.e. before and after vaccination). The cumulative cases since vaccination is a function of many variables, including timing of vaccination relative to the epidemic, force of infection, and VE values.

**Figure 4.**
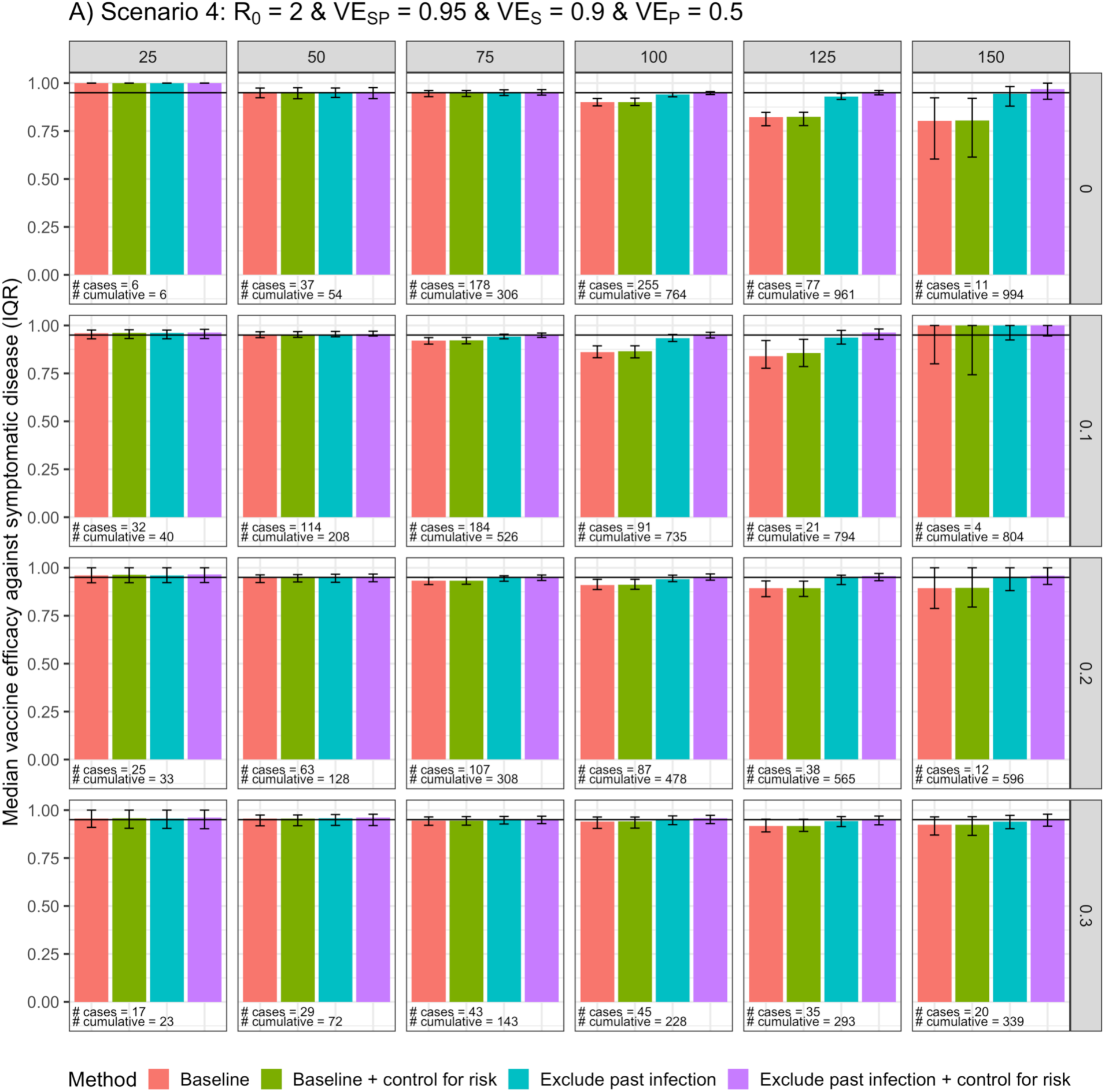
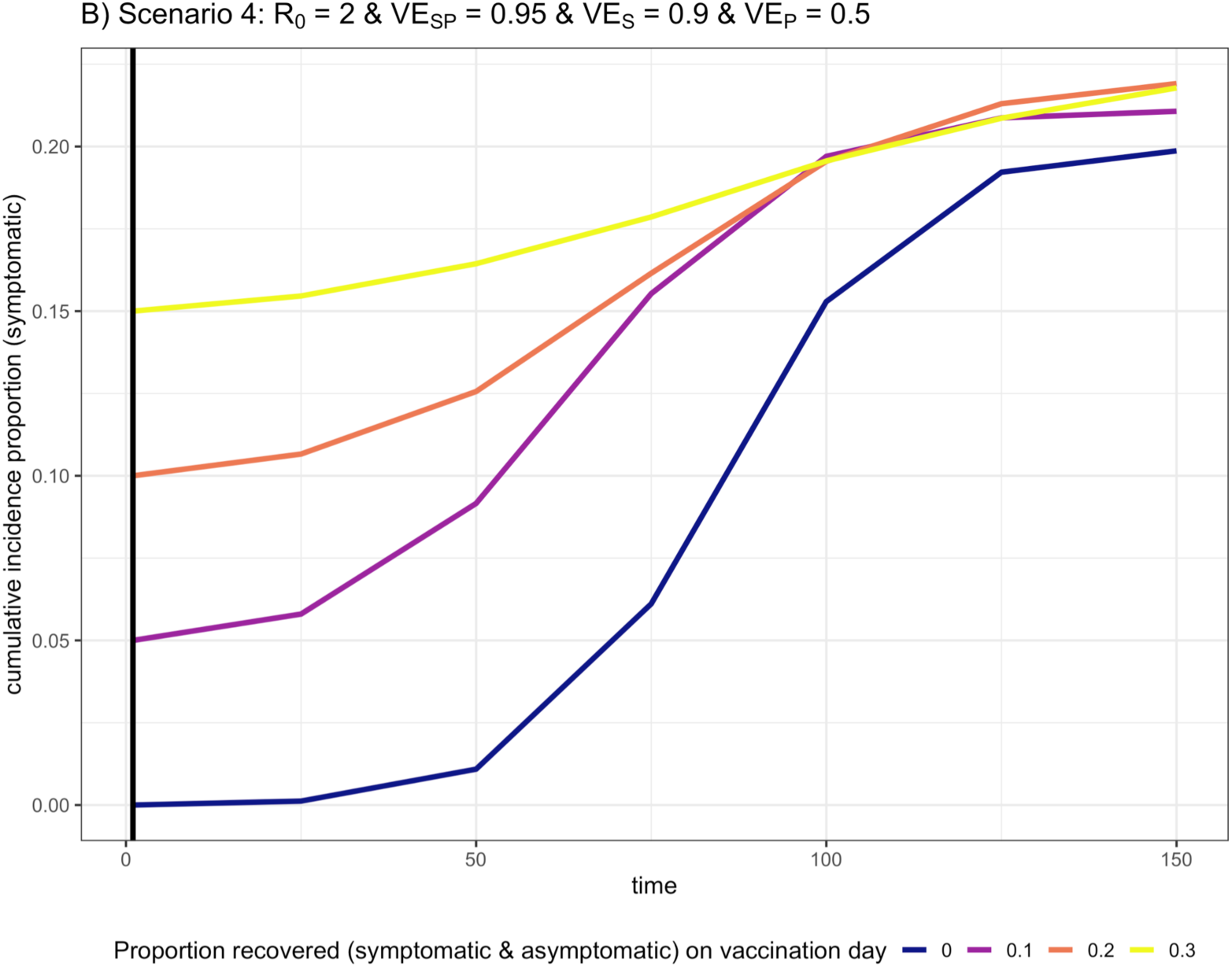
A) Vaccine efficacy against symptomatic disease for scenario 4 with a test-negative design. Columns are days since vaccination, and rows are the proportion infected before vaccination. Median and IQR of 100 simulations shown. Number of cases refers to the number of people with COVID-19 included in that day’s analysis. Cumulative number refers to the total number of cases of COVID-19 by that day since vaccination (denominator 5000). B) Median cumulative incidence proportion (symptomatic infection) over time since vaccination for the four values of the proportion recovered on vaccination day (black vertical line). Note the probability of symptoms for unvaccinated individuals is 50%.

In the cohort study analysis, which under our assumptions is equivalent to an RCT, we find similar trends to those observed in the TND (Figures 5-7); however, the cohort studies that do not exclude those with prior infection are less biased than equivalent TND studies for each scenario. In scenario 1, with the highest VESP, the bias is negligible. The bias is smaller because in cohort studies and RCTs, those with past symptomatic infection are censored at the time of infection, meaning fewer people are incorrectly treated as still at risk in the analysis.

**Figure 5.**
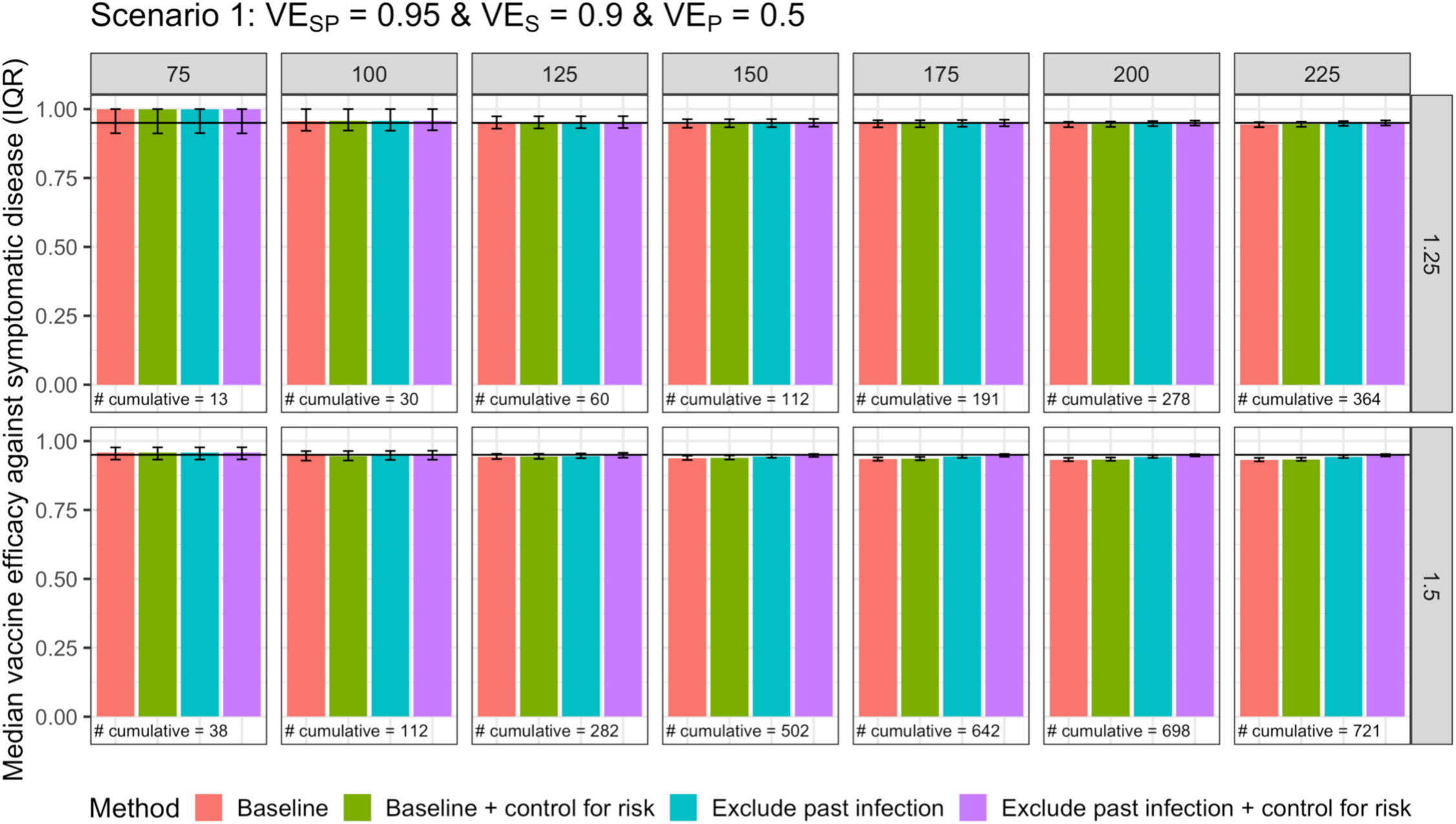
Vaccine efficacy against symptomatic disease for scenario 1 with a cohort/RCT study design. Columns are days since vaccination, and rows are values of R0. Median and IQR of 100 simulations shown. Cumulative number refers to the median total number of cases of COVID-19 by that day since vaccination (denominator 5000).

**Figure 6.**
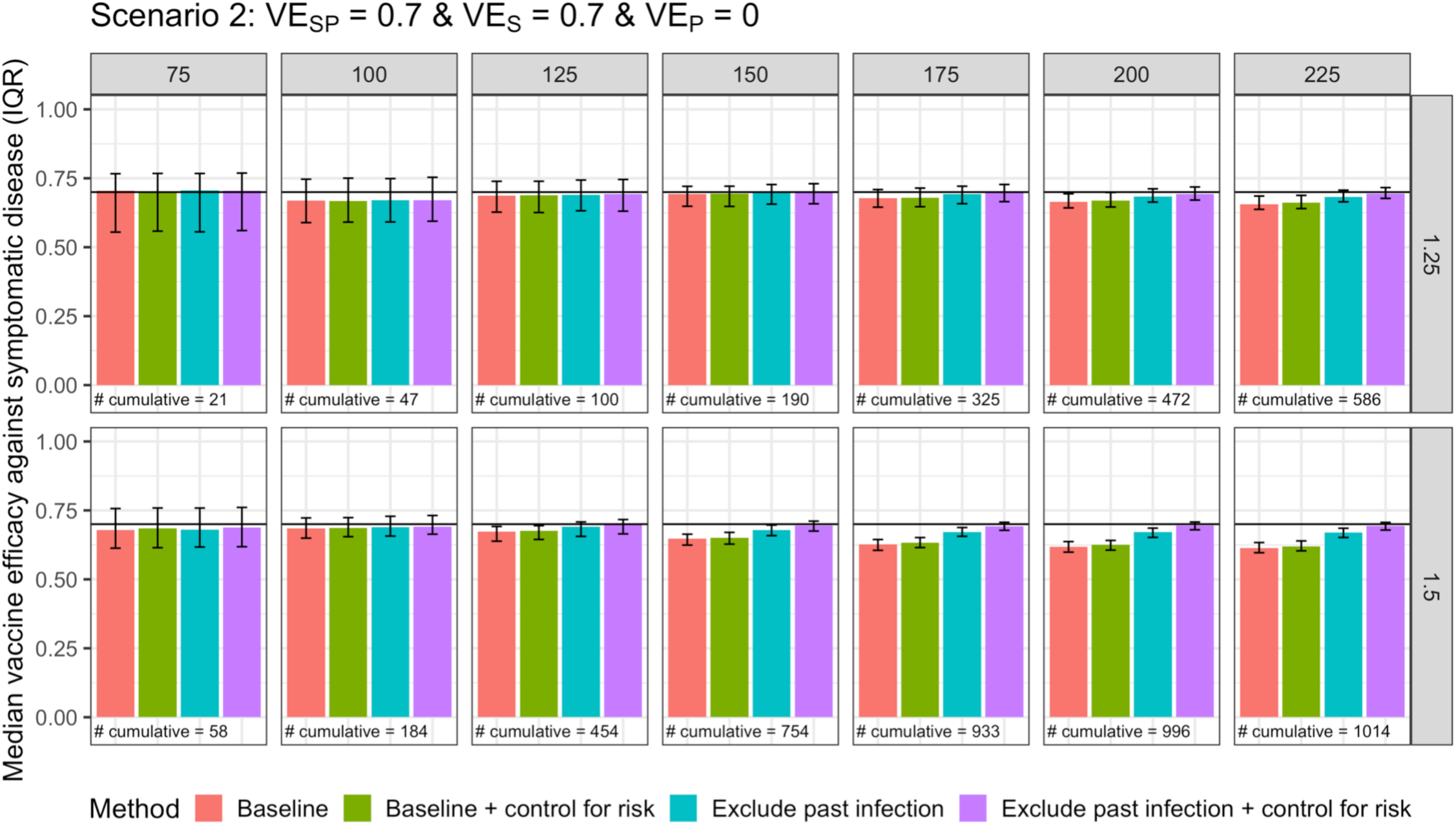
Vaccine efficacy against symptomatic disease for scenario 2 with a cohort/RCT study design. Columns are days since vaccination, and rows are values of R0. Median and IQR of 100 simulations shown. Cumulative number refers to the median total number of cases of COVID-19 by that day since vaccination (denominator 5000).

**Figure 7.**
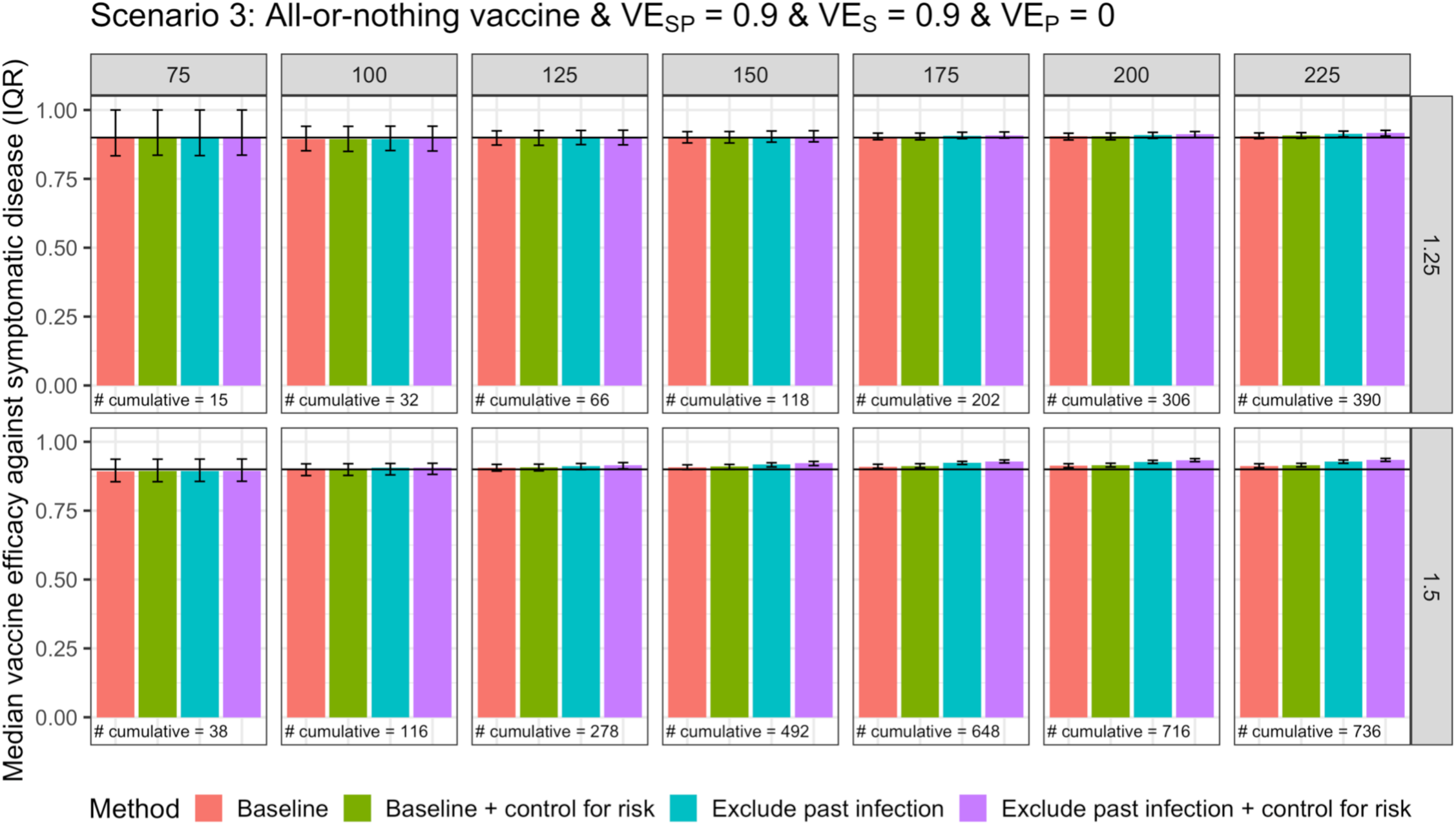
Vaccine efficacy against symptomatic disease for scenario 3 with a cohort study design. Columns are days since vaccination, and rows are values of R0. Median and IQR of 100 simulations shown. Cumulative number refers to the median total number of cases of COVID-19 by that day since vaccination (denominator 5000).

In sensitivity analyses, we relax assumptions of perfect tests for prior infection and examine lower sensitivity for both cases and controls and lower specificity for cases. We find that while lower sensitivity results in slight downward biases of the estimates, which are more pronounced in scenario 2 than scenario 1 (Figures S3-S4), lower specificity for cases does not induce a bias (Figure S5). This is because imperfect specificity only reduces the sample size, but we assume it does not do so differentially by vaccination status. A study of breakthrough infections in Israel found antibody levels on day of diagnosis were not greatly impacted by the current infection, suggesting imperfect specificity may not be a large concern (25).

Finally, in analyses that vary the parameters for reduction in susceptibility following infection and the proportion of the population at high risk, we find similar results to the baseline scenario (Figures S6-7). In analyses with a higher proportion symptomatic, we find less bias as expected given that a smaller proportion of cases will go undetected (Figure S8); we focus here on the cohort/RCT designs in which all symptomatic cases are identified and therefore the proportion symptomatic is a key parameter of interest.

## Discussion

We find that in scenarios with high baseline VE, differential depletion of susceptibles creates minimal bias in VE estimates and in the time trend of these estimates; therefore, there is little suggestion of spurious waning from comparing later to earlier VE estimates. While it is important to control for known predictors of risk, estimates that do not account for prior infection status will likely not be far off from the truth. In fact, without knowledge of the vaccine mechanism (i.e. leaky or all-or-nothing), it may be better to not condition on prior infection status: if the vaccine is leaky, the baseline estimates may be slightly underestimated, but if the vaccine is all-or-nothing, the adjusted estimates will overestimate the true VE. This upward bias occurs because excluding people with past infection with an all-or-nothing vaccine removes people for whom the vaccine did not work at all and focuses the analysis on those for whom the vaccine may be effective; with leaky vaccines, the individuals who are removed in the adjusted analysis are random (after accounting for risk factors). Evaluating how the estimates from different analyses change over time could give potential insight into the type of vaccine mechanism.

Because the bias from failing to exclude prior infection in the analysis of a leaky vaccine with high initial VE is expected to be small under the null hypothesis of no waning VE, if the VE appears to wane substantially, this finding is likely not entirely due to bias. If true waning occurs, spurious waning bias may become a more relevant consideration (as in scenarios with a lower baseline VE); that is, estimates may reflect a combination of real and spurious waning. Six month efficacy results from the mRNA vaccine trials show mixed findings regarding waning, with Moderna showing consistent efficacy over time (26) and Pfizer’s estimates slightly declining (27). It is challenging to disentangle if this decline is due to lower effectiveness against variants, true waning, spurious waning, or some combination of these factors; given the minimal bias found in our RCT-like analysis of vaccines with high VE (Figure 5), our findings suggest the decline is likely not due to spurious waning alone. Similarly, spurious waning is likely not the only cause of the declines in effectiveness observed in Israel, given the high effectiveness estimated when vaccines were first given (19,20), the magnitude of the declines and that they occurred following a period of low incidence (28).

If baseline VE is lower, the bias over time for leaky vaccines is larger and ideally should be corrected if the mechanism is known to be leaky. However, leaky and all-or-nothing mechanisms are two extremes; in reality, vaccines will fail to take in some individuals due to improper handling or injection so most vaccines are leaky vs. nothing. By examining both mechanisms, our analyses show the range of possible biases. In the absence of other sources of bias, conducting analyses both unadjusted and adjusted for past infection could give lower and upper bounds for the true VE. Studies of VE over time should therefore enroll individuals regardless of prior infection history but also collect information on this critical variable for use in the analysis; when possible, prior infection status should be assessed using serology as even an imperfect serologic test will improve sensitivity over self-report alone.

This study has several limitations. First, we make many simplifying assumptions in the model. For example, we assume all individuals are grouped into one large community and do not examine the potential impact of geographic heterogeneity. Other studies have shown epidemic dynamics due to differences in geography are important to control for in vaccine (29) and serologic (5) studies. We also assume perfect sensitivity and specificity of virologic tests, as implications of these parameters have been explored in detail previously (30,31). There are many potential biases in studies of vaccine effectiveness, which are described in detail in World Health Organization guidance (30); here we focus specifically on spurious waning bias from differential depletion of susceptibles. While we incorporate heterogeneity in risk of acquiring infection, we do not model differences in risk of transmitting infection (e.g. due to host factors). Second, using serologic tests to identify prior infection is subject to error from imperfect test characteristics and waning of antibodies over time. However, we find only small biases in VE estimates from imperfect sensitivity, and information on past infection can also be obtained through self-report or medical records. Third, as described above, we assume random vaccination and no unmeasured confounding; the strategies discussed here alone do not address most other sources of potential confounding, which are important to account for in analyses, particularly given that vaccine rollout in some cases prioritized those at highest risk to receive vaccines first. Fourth, we simulated epidemics with higher R values than much of the United States experienced during most of the pandemic to uncover scenarios where spurious waning might be of concern (32). These values should not affect the conclusions from the simulations, as we find that the main determinant of the extent and magnitude of bias is the proportion of the population that has been infected since vaccination, which is influenced by a combination of several factors, including R values, the point in the epidemic trajectory when the vaccine was introduced, and prevalence of high vs low risk individuals in the population. Finally, we assume no true waning or other reasons for decreased effectiveness, such as new variants; future research should explore methods for disentangling these potential explanations for observed declines in effectiveness over time.

Assessing duration of protection from COVID-19 vaccines is important for anticipating future dynamics of this pandemic. Here we have outlined circumstances under which bias can arise in these estimates and identified approaches to alleviate these biases.

## Data Availability

Code is available on github.

https://github.com/rek160/spurious-waning

## Acknowledgments

We thank Rachel Slayton and Matt Biggerstaff for helpful comments and discussion.

## Funding

This work was supported by the U.S. National Cancer Institute Seronet cooperative agreement U01CA261277. The content is solely the responsibility of the authors and does not necessarily represent the official views of the National Institutes of Health.

## Conflicts of interest

Dr. Lipsitch reports consulting/honoraria from Bristol Myers Squibb, Sanofi Pasteur, and Merck, as well as a grant through his institution, unrelated to COVID-19, from Pfizer. He has served as an unpaid advisor related to COVID-19 to Pfizer, One Day Sooner, Astra-Zeneca, Janssen, and COVAX (United Biomedical). Dr. Kahn discloses consulting fees from Partners In Health.

---

**Figure S1.**
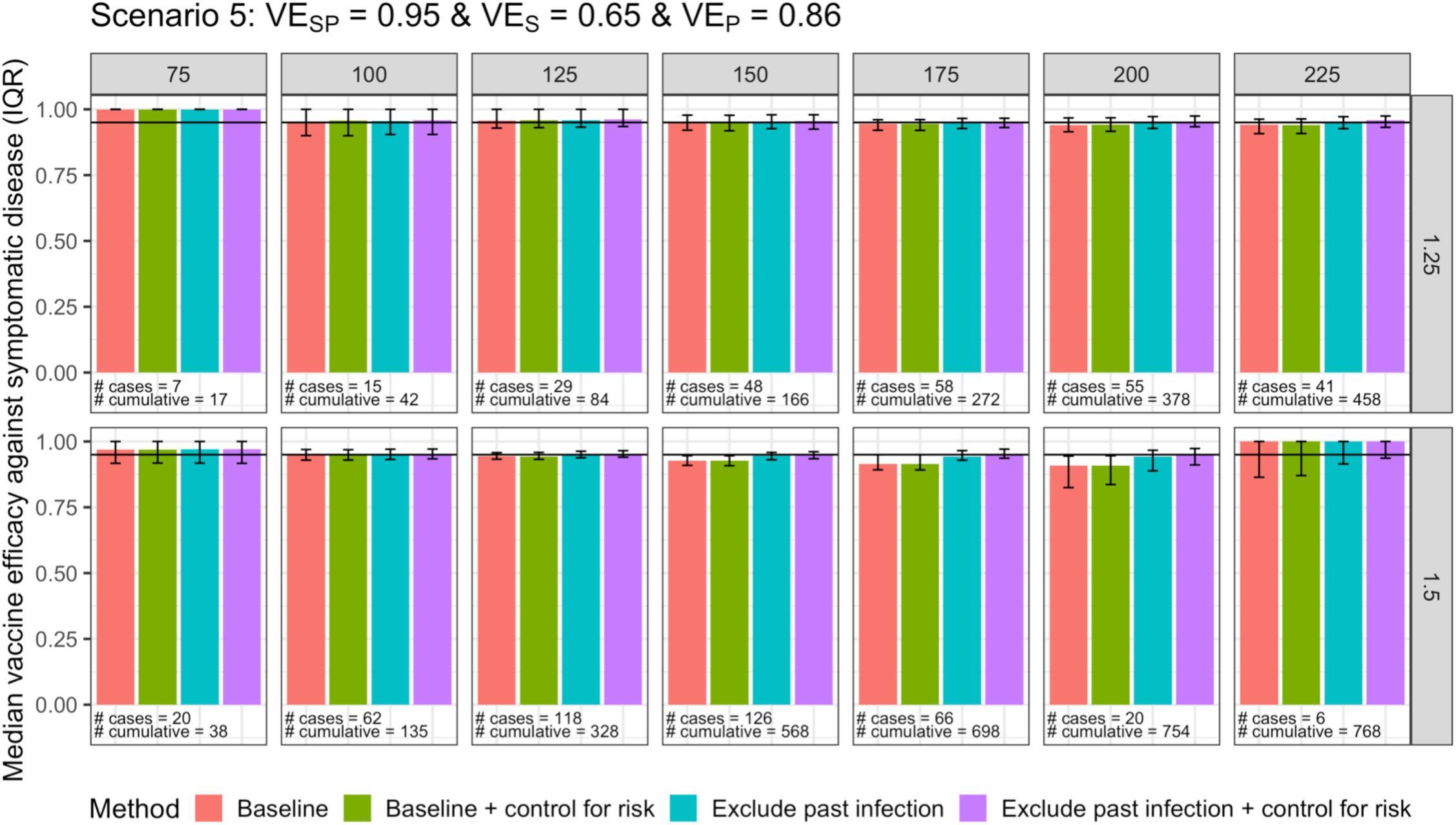
Vaccine efficacy against symptomatic disease for scenario 5 with a test-negative design. Columns are days since vaccination, and rows are values of R. Median and IQR of 100 simulations shown. Number of cases refers to the median number of people with COVID-19 included in that day’s analysis. Cumulative number refers to the median total number of cases of COVID-19 by that day since vaccination (denominator 5000).

**Figure S2.**
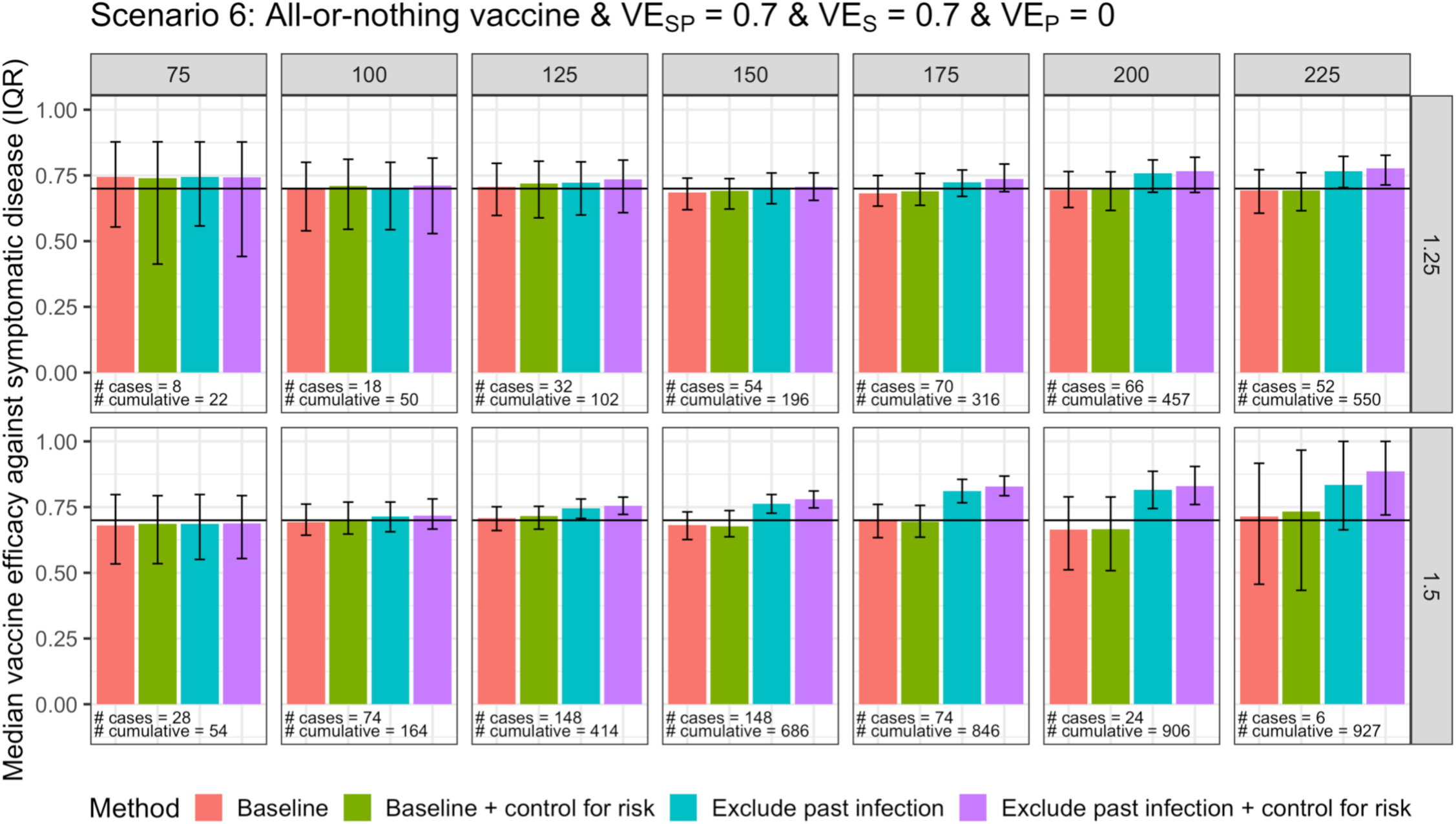
Vaccine efficacy against symptomatic disease for scenario 6 with a test-negative design. Columns are days since vaccination, and rows are values of R. Median and IQR of 100 simulations shown. Number of cases refers to the median number of people with COVID-19 included in that day’s analysis. Cumulative number refers to the median total number of cases of COVID-19 by that day since vaccination (denominator 5000).

**Figure S3.**
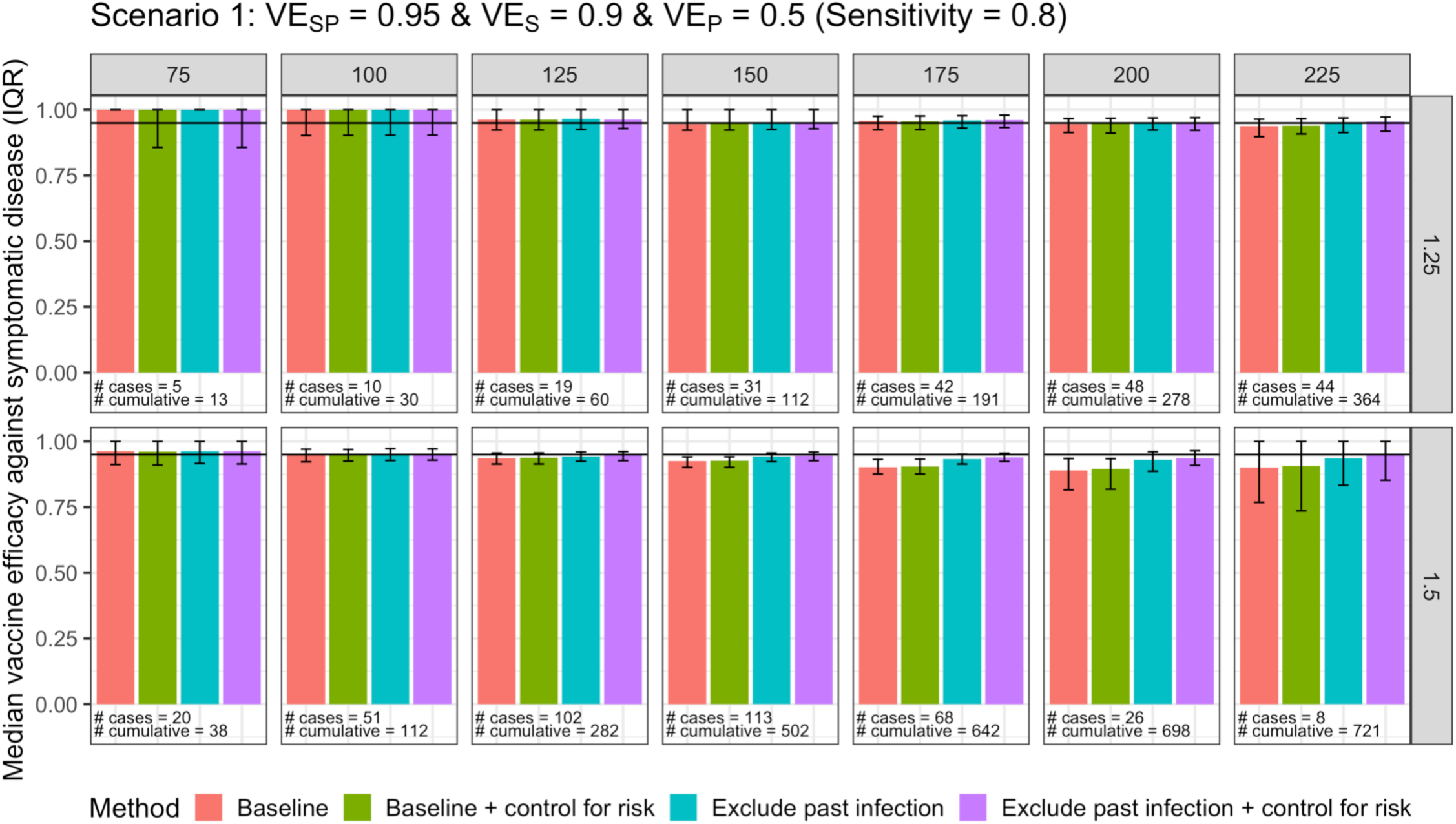
Vaccine efficacy against symptomatic disease for scenario 1 with imperfect sensitivity for the test for prior infection with a test-negative design. Columns are days since vaccination, and rows are values of R. Median and IQR of 100 simulations shown. Number of cases refers to the median number of people with COVID-19 included in that day’s analysis. Cumulative number refers to the median total number of cases of COVID-19 by that day since vaccination (denominator 5000).

**Figure S4.**
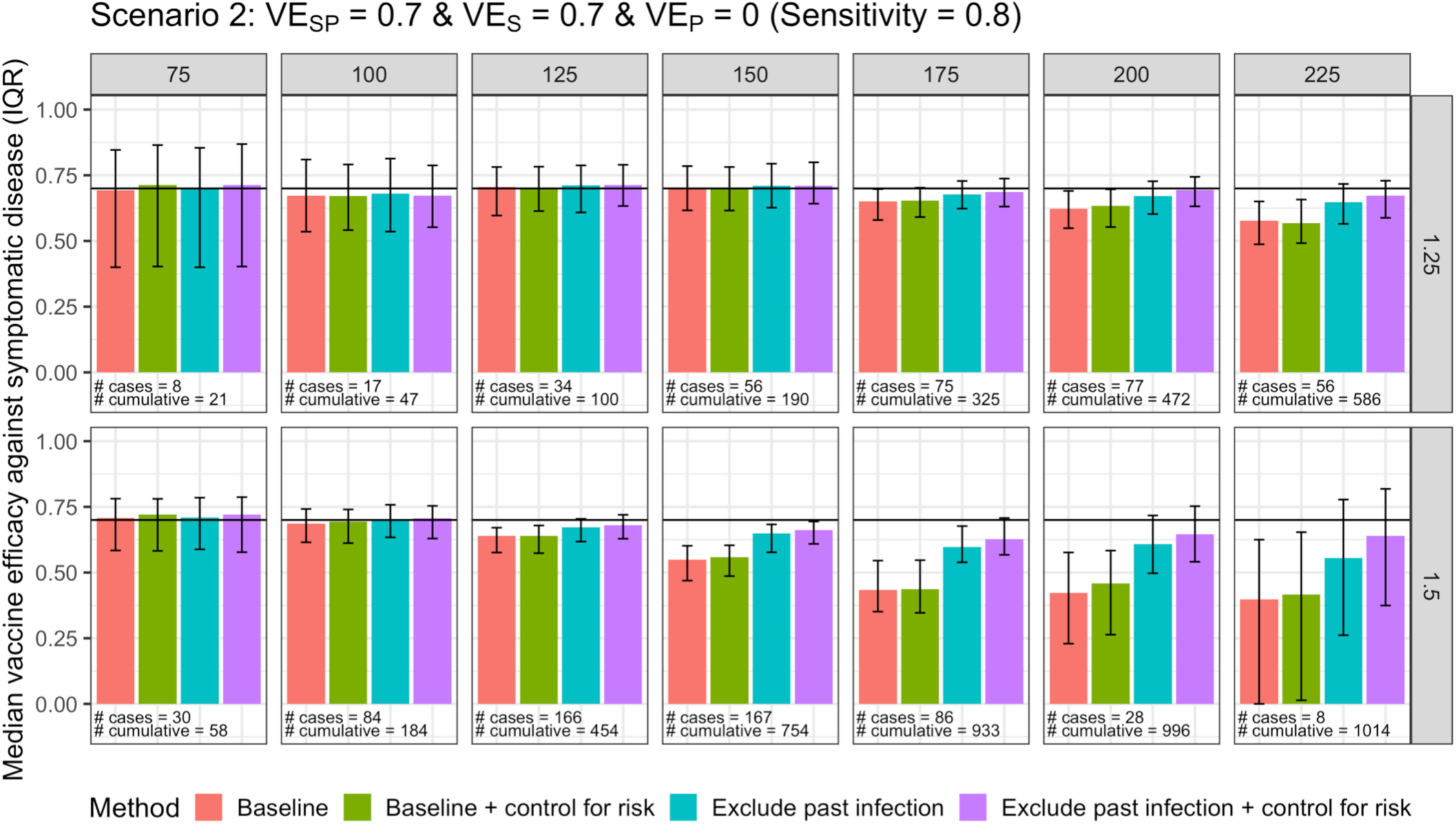
Vaccine efficacy against symptomatic disease for scenario 2 with imperfect sensitivity for the test for prior infection with a test-negative design. Columns are days since vaccination, and rows are values of R. Median and IQR of 100 simulations shown. Number of cases refers to the median number of people with COVID-19 included in that day’s analysis. Cumulative number refers to the median total number of cases of COVID-19 by that day since vaccination (denominator 5000).

**Figure S5.**
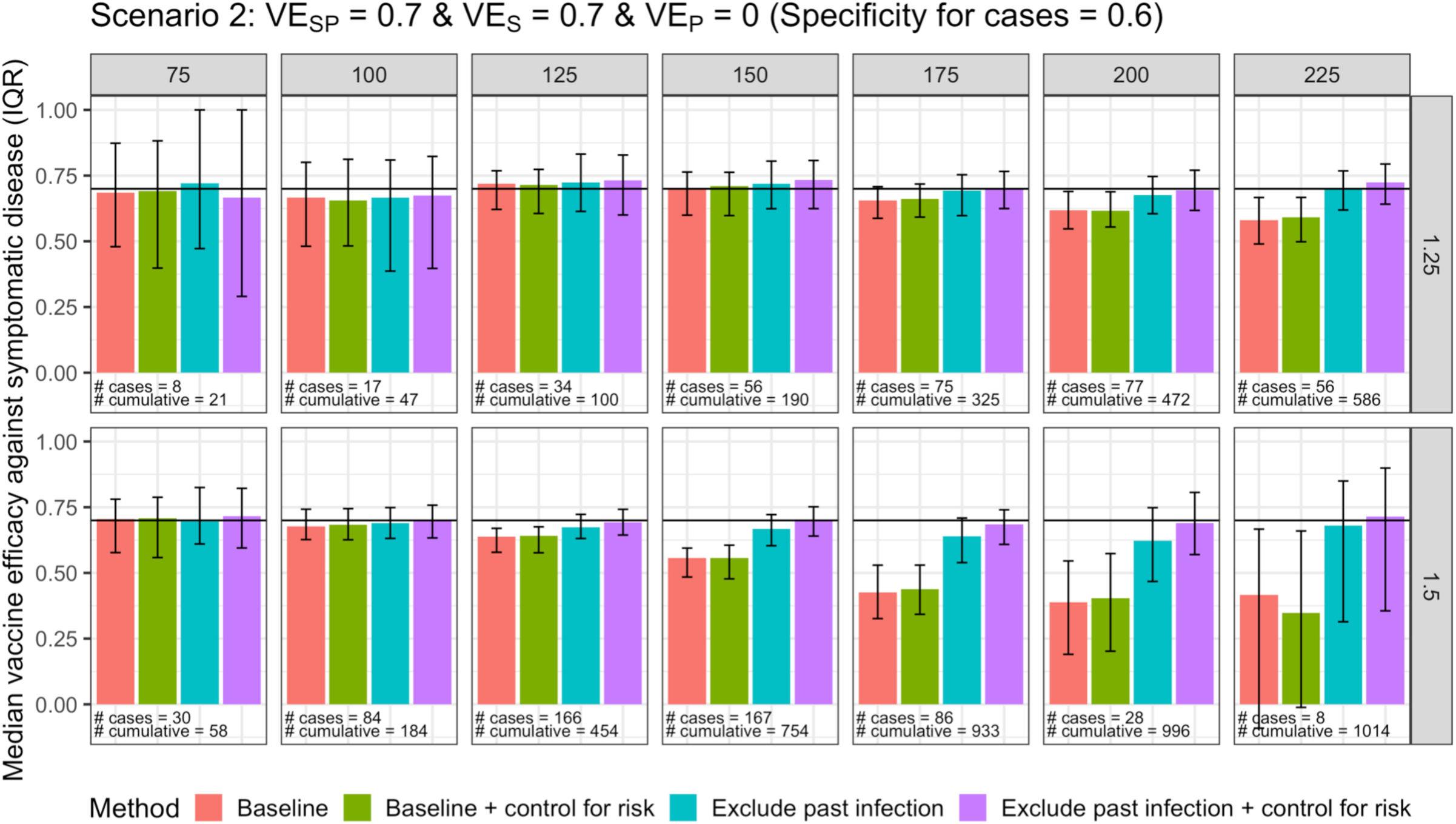
Vaccine efficacy against symptomatic disease for scenario 2 with imperfect specificity for cases for the test for prior infection with a test-negative design. Columns are days since vaccination, and rows are values of R. Median and IQR of 100 simulations shown. Number of cases refers to the median number of people with COVID-19 included in that day’s analysis. Cumulative number refers to the median total number of cases of COVID-19 by that day since vaccination (denominator 5000).

**Figure S6.**
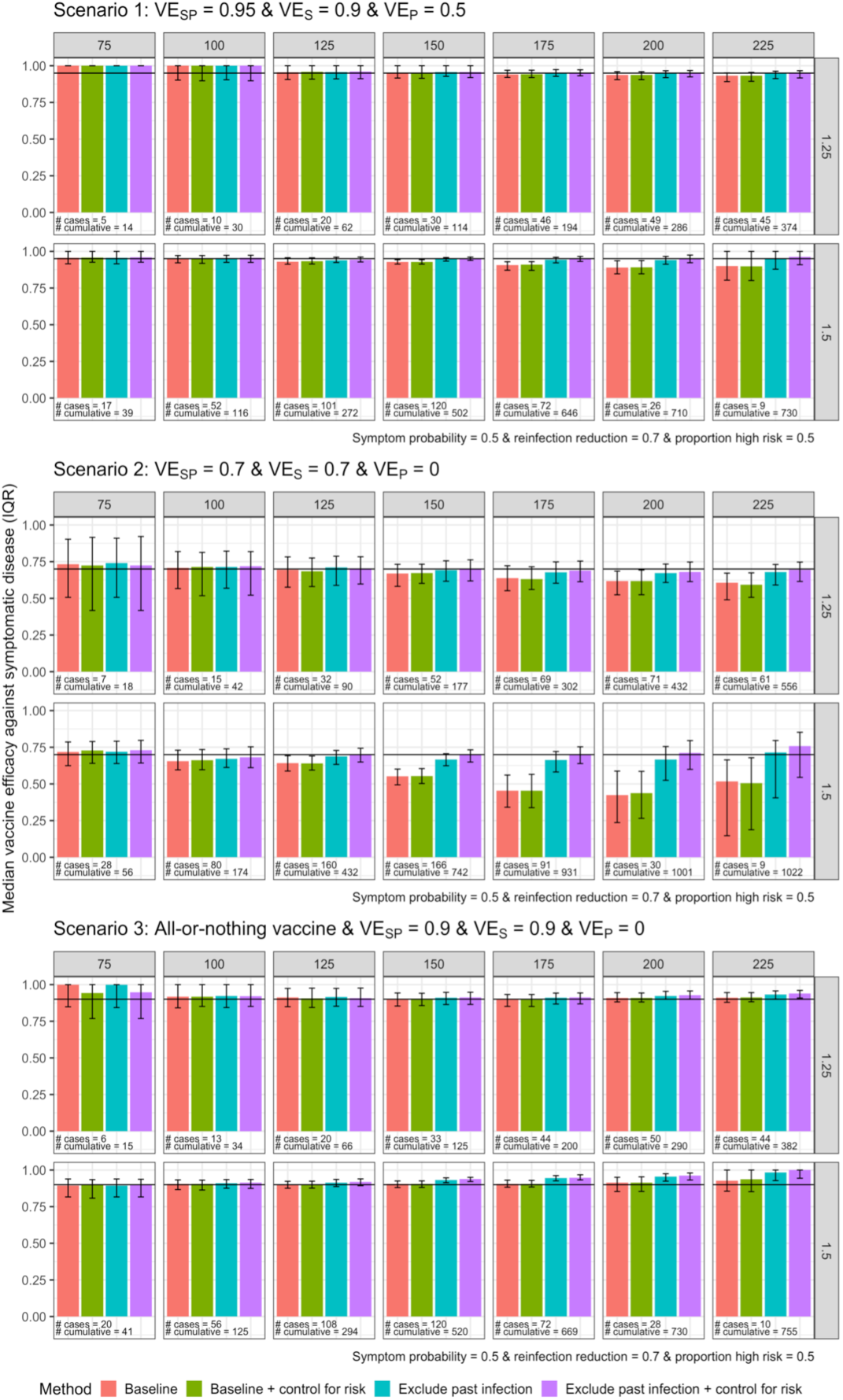
Vaccine efficacy against symptomatic disease for scenarios 1-3 with a test-negative design, with lower relative reduction in susceptibility following infection compared to baseline parameters. Columns are days since vaccination, and rows are values of R. Median and IQR of 100 simulations shown. Number of cases refers to the median number of people with COVID-19 included in that day’s analysis. Cumulative number refers to the median total number of cases of COVID-19 by that day since vaccination (denominator 5000).

**Figure S7.**
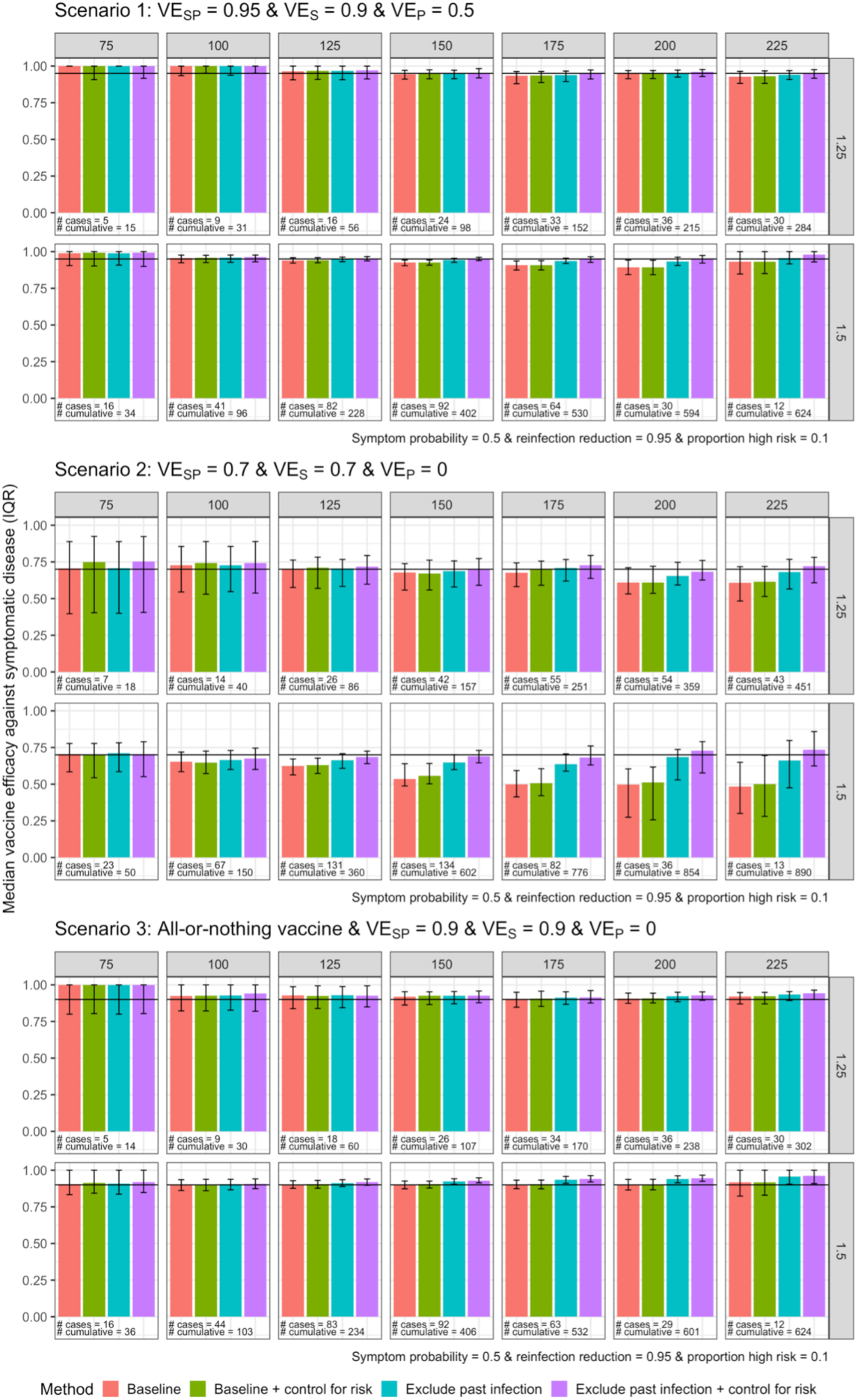
Vaccine efficacy against symptomatic disease for scenarios 1-3 with a test-negative design, with lower proportion of the population at high risk compared to baseline parameters. Columns are days since vaccination, and rows are values of R. Median and IQR of 100 simulations shown. Number of cases refers to the median number of people with COVID-19 included in that day’s analysis. Cumulative number refers to the median total number of cases of COVID-19 by that day since vaccination (denominator 5000).

**Figure S8.**
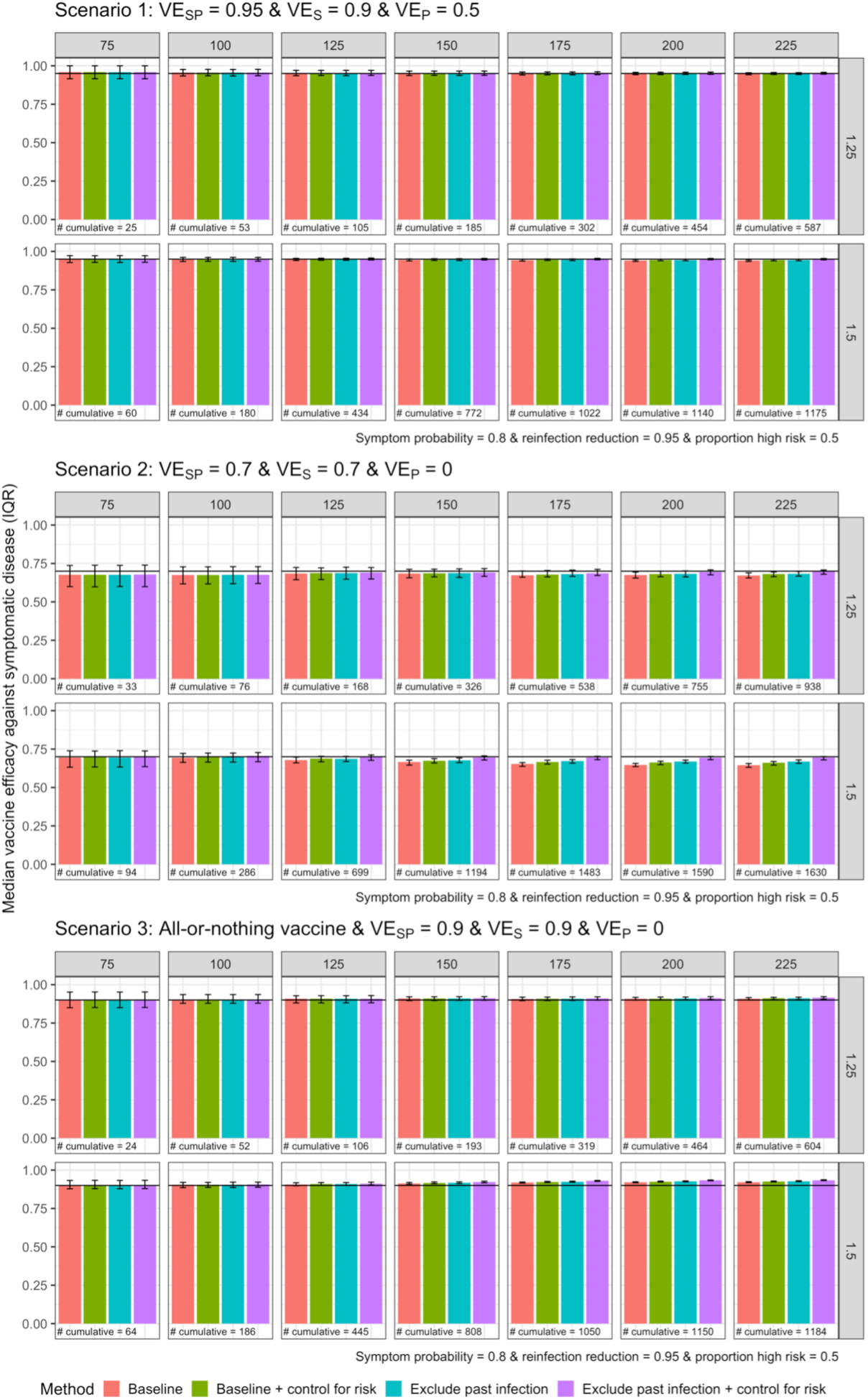
Vaccine efficacy against symptomatic disease for scenarios 1-3 with a cohort/RCT design, with higher proportion symptomatic compared to baseline parameters. Columns are days since vaccination, and rows are values of R. Median and IQR of 100 simulations shown. Number of cases refers to the median number of people with COVID-19 included in that day’s analysis. Cumulative number refers to the median total number of cases of COVID-19 by that day since vaccination (denominator 5000).

